# Preterm birth and the risk of infant respiratory syncytial virus associated acute lower respiratory infection in Australia’s Northern Territory

**DOI:** 10.1101/2025.06.30.25330542

**Authors:** Michael J Binks, Tejal Shah, Sasha Underhill, Jemima Beissbarth, Robyn Marsh, Lisa McHugh, Peter S Morris, Kiarna Brown, Bianca Middleton

**Author notes:** **Address correspondence to:** Michael J Binks, John Matthews Building (58), Menzies School of Health Research, Royal Darwin Hospital Campus, Tiwi, Northern Territory, 0811. **Author contributions** Associate Professor Michael Binks conceived the project, led the data extraction, data linkage, cleaning, analysis, interpretation and prepared the article. Dr Bianca Middleton provided RSV specific and general paediatric clinical knowledge and interpretation. Dr Kiarna Brown provided obstetric and general clinical interpretation. Professor Peter Morris provided design oversight, clinical interpretation and expert statistical advice. All authors contributed intellectual input to the design, and preparation of the Article, and approved the submission for publication.

## Abstract

**Objective:** The Northern Territory (NT) of Australia has high rates of both preterm birth and childhood respiratory syncytial virus (RSV) infections. This study evaluated preterm birth and subsequent risk of RSV-associated acute lower respiratory infection hospital admission (RSV-ARLI) among NT infants during their first year of life.

**Methods:** A retrospective, population-based cohort study of NT mother-infant pairs from 2008 to 2017. Preterm birth was defined as gestation <37 weeks. RSV-ALRI hospitalisations were identified using ICD-10-AM codes (J12.1, J20.5, J21.0, or J09-J22 plus B97.4). Risk of RSV-ALRI in preterm infants was assessed using generalised linear regression to calculate risk ratios (RR). Two multivariable models were used: one including all covariates (aRR1) and another with stepwise elimination retaining only significant factors (aRR2).

**Results:** Overall, 10% (3839/39148) of infants were born preterm and 2% (846/39148) were hospitalised with RSV-ALRI. Preterm infants (4%) had a significantly higher risk or RSV-ALRI than term infants (2%) (risk ratio [RR] 2.26, 95% CI 1.90-2.69). This persisted even after adjustment for demographic, antenatal and infant health characteristics (aRR1 1.72, 1.43-2.08; aRR2 1.83, 95% CI 1.52-2.19). The risk of RSV-ALRI was highest among infants born <31 weeks gestation (7%). First Nations infants (4%) and those from very remote central desert regions (7%) also had a high risk of RSV-ALRI hospitalisation.

**Conclusion:** Preterm birth is a common and significant risk factor for infant RSV hospitalisation in Australia’s NT. RSV immunisation strategies will be crucial in protecting high-risk infants in the region.

**Article Summary:** Preterm birth doubles RSV hospitalisation risk in Australia’s Northern Territory. First Nations children and those in remote regions experienced the highest RSV burden.

**What’s Known on This Subject:** Preterm birth and acute respiratory infections contribute substantially to infant morbidity in Australia’s Northern Territory. Globally, respiratory syncytial virus is a leading cause of hospitalisation, and preterm infants account for a disproportionate share of RSV-associated admissions in early childhood.

**What This Study Adds:** This is the first population-based study quantifying the RSV hospitalisation risk among preterm infants in northern Australia. Preterm birth significantly increases RSV hospitalisation risk. First Nations children and remote populations were identified as priority prevention targets.

## Introduction

Preterm birth is a significant concern in Australia’s Northern Territory (NT), with a reported prevalence as high as 16% among First Nations families. ^1^ NT infants also have among the world’s highest rates of acute lower respiratory infection (ALRI) requiring hospitalisation (20% in the first year of life). Respiratory syncytial virus is a leading respiratory pathogen. ^2^ As many as 8% of First Nations children living in remote NT communities are hospitalised with RSV-associated ALRI (RSV-ALRI) in their first year of life, ^3^ causing significant social disruption and strain on health services. There is evidence to suggest that RSV seasonality is unique in the NT and varies by region, correlating with high rainfall and humidity (December to May) in the tropical Top End^3,4^ and with winter/spring seasons (May to September) in the temperate Central Desert region. ^3^

Globally, preterm birth is a prominent risk factor for early childhood RSV infections. A 2024 meta-analysis of 47 studies found preterm infants account for 25% of all RSV-ALRI hospitalisations in children under two years of age. ^2^ Another review of 20 studies found preterm infants have nearly double the risk of RSV-ALRI hospitalisation (odds ratio [OR] 1.96, 95% CI 1.44–2.67), with additional important risk factors including low birth weight, maternal smoking, crowded households, male sex, and lack of breastfeeding. ^5^ Preterm birth is also linked to severe RSV outcomes and increased infant mortality. ^6^ Few Australian data exist, but a New South Wales study found preterm infants had a 2.5-fold higher incidence of RSV-coded hospitalisations during the first six months of life. ^7^

There are no published data on the association between preterm birth and RSV-ALRI hospitalisation in the NT, no longitudinal data on preterm birth in the Central Desert region and no published NT RSV-ALRI incidence data since 2015. This study evaluates the association between preterm birth and RSV-ALRI hospital admissions among NT infants (2008–2017), to guide prevention strategies including maternal RSV vaccination and infant immunoprophylaxis.

## Methods

### Study design, population and setting

This retrospective population-based cohort study included all infants born in the NT from January 2008 to December 2017 (Figure 1). The NT covers 1.4 million km,² with 233,000 residents (2021), 26% identifying as First Nations, and most living outside Greater Darwin. ^8,9^ Five public hospitals and one small private hospital serviced the region, with acute paediatric inpatient care managed almost exclusively by the public hospitals.

**Figure 1.**
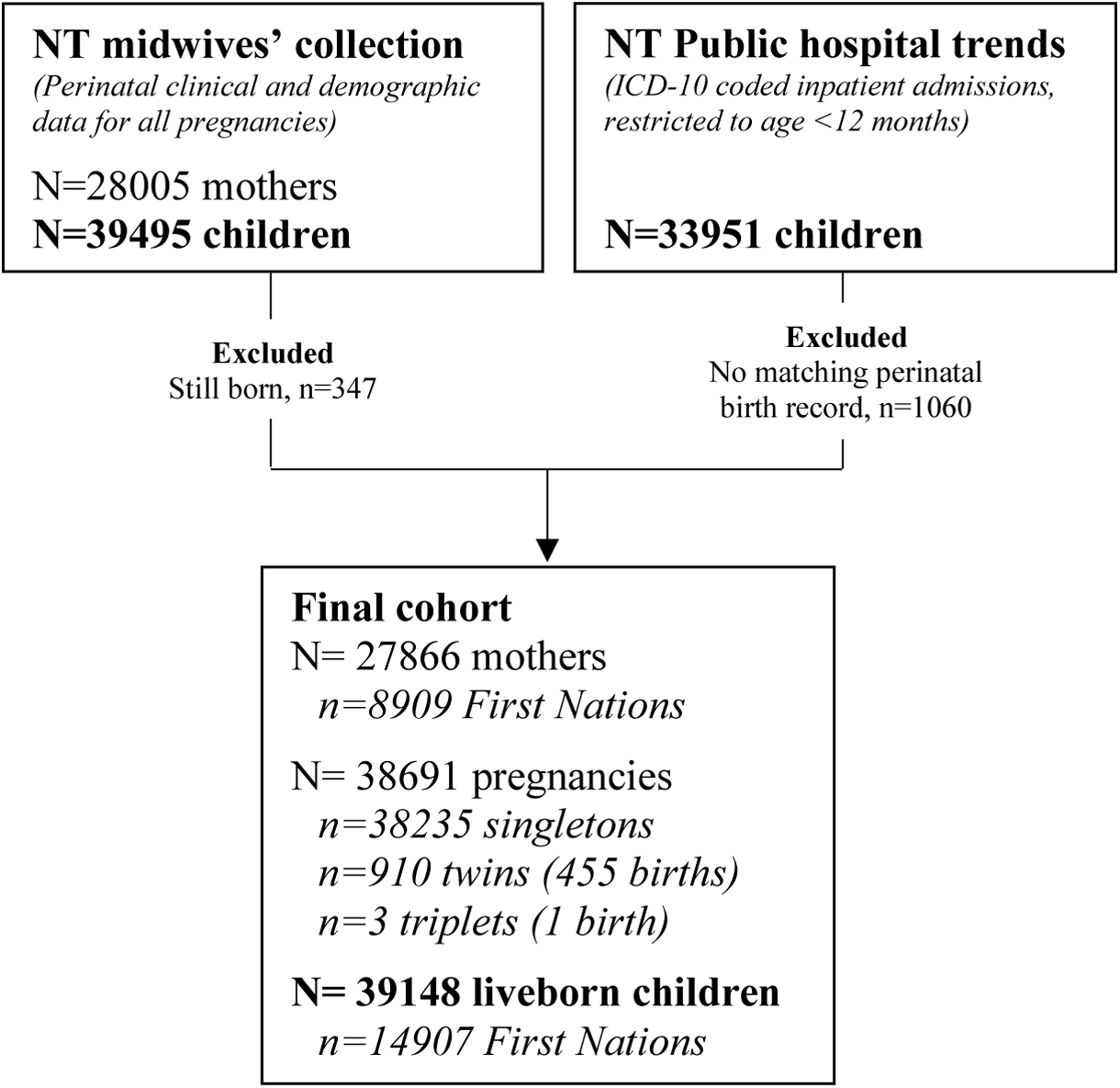
Northern Territory (NT) birth cohort: 2008-2017. The cohort was defined by a live-birth record in the NT perinatal dataset during the study period. Hospital admissions for children within their first 12 months of life, identified through ICD-10 coding records in the NT public hospital trends dataset, were deterministically linked to their corresponding perinatal birth records using unique identifiers. Children who were not hospitalised in their first 12 months were assumed to be NT residents with no hospital admissions during infancy. Hospital admissions occurring outside the NT are a potential source of under-ascertainment. Children who were hospitalised but could not be matched to a perinatal birth record were presumed to have been born outside the NT and were excluded from the analysis. During the study period, all acute paediatric hospital admissions in the NT were serviced by five public hospitals: Royal Darwin Hospital, Katherine District Hospital, Gove District Hospital, Tennant Creek Hospital, and Alice Springs Hospital. The NT’s sole private hospital offered birthing services but provided almost no acute paediatric in-patient care.

### Data sources and linkage

Data were sourced from the NT Midwives’ Collection (perinatal data) and NT Territory Hospital Trends Inpatient Activity (hospital admissions). The Midwives’ Collection includes all births ≥20 weeks or ≥400g or any signs of life at birth, with gestational age based on last menstrual period or earliest ultrasound. First Nations status was validated against master index and hospital records. ^10^ The Hospital Trends dataset contains ICD-10-AM coded summaries for all public hospital admissions. Datasets were linked deterministically using a project-specific key.

### Exposure and outcomes

Exposure: preterm birth, was defined as <37 completed weeks gestation, with sub classifications 34-36, 32-33, 28-31 and 20-27 weeks. ^11,12^

Primary outcome: RSV-ALRI hospitalisation defined by the ICD-10-AM codes: J12·1 or J20.5 or J210 or J09-J22 plus B97·4 (Supplementary Figure 1). Repeat admissions within 7 days were consolidated into a single episode. All diagnostic codes recorded for an admission episode (primary, secondary, tertiary) contributed to outcome classification. That is, RSV-ARLI hospitalisation did not need to be the primary diagnosis associated with hospital admission and a single hospital admission could have more than one respiratory outcome classification.

Other outcomes: Viral and bacterial pneumonia, bronchitis, bronchiolitis, and non-specific ALRI (Supplementary Figure 1). Non-respiratory conditions, gastroenteritis (A00-A09.9), skin infection (L00-L08.9, or P39.4) and scabies (B86) were analysed for specificity.

### Analysis

All analyses were conducted using Stata software version 17.

### Descriptive analysis

Antenatal, delivery and infant characteristics were stratified by the preterm birth exposure. Between group differences were assessed using univariate risk ratio (RR) with 95% confidence interval (95% CI).

### Geography classifications

Geography was categorised using Local Government Areas (Top End, Central Desert) and ARIA+ Remoteness Areas (outer regional, remote, very remote). See Supplementary Methods, page 13, for full details.

### Preterm birth and RSV-ALRI hospitalisation risk was evaluated using

1. **Univariable analysis**: Crude risk ratios (RRs) with 95% confidence intervals (95% CIs) were calculated to estimate the unadjusted risk of RSV-ALRI hospitalisation in preterm infants relative to term-born infants.
2. **Multivariable analysis**: Adjusted RRs (aRRs) were calculated using generalised linear models (log-binomial with robust error and clustering on family unit) accounting for potential confounders. Model 1 (Full Model, aRR1): adjusted for all available general maternal characteristics, pre-existing health risks, obstetric complications, and birth/infant characteristics (as per Table 2). Model 2 (Parsimonious model, aRR2): used backward elimination (p<0.05) to identify the most robust predictors of RSV-ALRI hospitalisation risk. Both models were adjusted for secular (year of birth) and seasonal (winter/spring/summer/autumn) trends. Robust standard errors clustered by family were used to address sibling correlations. Missing data were retained as a separate unknown category. Goodness-of-fit was assessed using pseudo R^2^ supported by Akaike Information Criterion (AIC) and Bayesian Information Criterion (BIC). RRs were considered statistically significant where the 95% CI’s excluded 1.

### Age-specific risk

Age associated outcome trends were assessed through cumulative proportion (by age in months) and normalised frequency distribution (monthly episodes/total episodes) plots of first hospital admissions.

### Temporal trends

Hospitalisation incidence rates were calculated by diving the number of episodes by the number of children and time observed (episodes per 100 child-years). Each child was assumed to have had complete follow-up from birth to age 12 months. Each child could contribute more than one admission to the estimated hospitalisation incidence rate.

Annual hospitalisation incidence rates were plotted graphically. Temporal and between-group incidence rate trends were assessed using negative binomial regression, with annual differences and between-group comparisons (adjusted for year) presented as crude incidence rate ratios (IRRs). The effects of truncation bias were avoided in this birth cohort analysis by omitting data from 2008. Children born in 2007 but hospitalised with an ALRI in 2008 were not part of the cohort because their antenatal data was not available. Geographical variation in seasonality was visualised by aggregating and plotting monthly RSV-ALRI hospital admissions for Top End and Central Desert region over the study period.

## Results

### Preterm birth and cohort characteristics

The cohort (Figure 1) included 39,148 infants (27,866 mothers), with 10% (n=2,822) born preterm (<37 weeks): 6.8% at 34–36 weeks, 1.3% at 32–33 weeks, 1.1% at 28–31 weeks, and 0.7% at 20-27 weeks (Table 1a). Preterm birth risk was stable across 2008-2017 (Table 1b) and over twice as high in First Nations (14.6%) versus non-First Nations (6.8%) families.

**Table 1.**
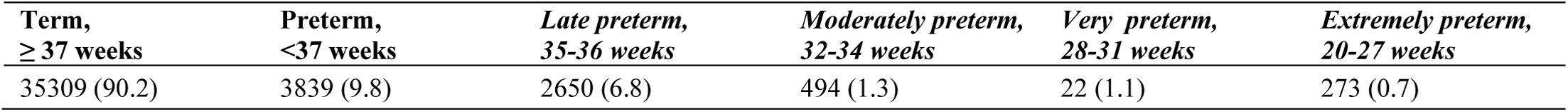
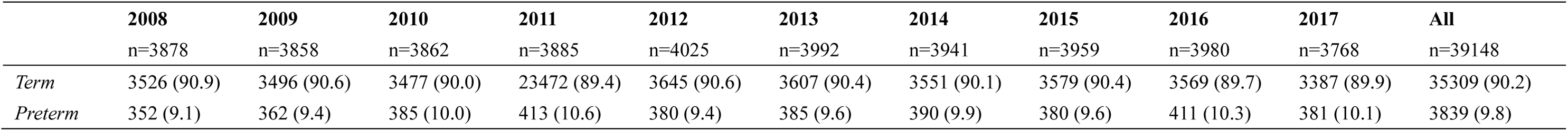
a. Overall prevalence of preterm birth and subcategories among Northern Territory infants, b. Prevalence of preterm birth among Northern Territory infants by year.

Seventy-five percent of pregnancies commenced antenatal care in the first trimester; 27.2% of pregnancies were among women living in very remote regions, and 23.9% involved maternal smoking. Gestational diabetes (10.7%) and anaemia (5.9%) were the most common complications. Most infants were born vaginally (68.5%), 17.6% were admitted to the special care nursery, and 38.1% were identified as Australian First Nations (Table 2).

**Table 2.** Characteristics of the mother-infant cohort in the Northern Territory (2008-2017): by preterm birth status.

|  | Overall children<br>(n=39148) | Preterm (<37 weeks) |  | Risk ratio (95% CI) |
| --- | --- | --- | --- | --- |
|  |  | Yes (n=3839, 9.8%) | No (n=35309, 90.2%) |  |
| **General characteristics, n(%)** |  |  |  |  |
| First Nations mother | 13670 (34.9) | 2095 (54.6) | 11575 (32.8) | 1.66 (1.61,1.72) |
| **Maternal age at birth (years)** |  |  |  |  |
| ≤19 | 3346 (8.6) | 427 (11.1) | 2919 (8.3) | 1.45 (1.29,1.62) |
| 20-29 | 18905 (48.3) | 1902 (49.5) | 17003 (48.2) | 1.14 (1.06,1.22) |
| 30-39 | 15720 (40.2) | 1385 (36.1) | 14335 (41.6) | ref |
| ≥40 | 1177 (3.0) | 125 (3.3) | 1052 (3.0) | 1.21 (0.99,1.46) |
| First pregnancy | 11919 (30.6) | 1108 (28.9) | 10811 (30.6) | 0.94 (0.89,0.99) |
| Antenatal care in 1<sup>st</sup> trimester | 29137/38736 (75.2) | 2540/3707 (68.5) | 26597/29137 (75.9) | 0.90 (0.88,0.92) |
| Any smoking during pregnancy | 8750/36592 (23.9) | 1209/3407 (35.5) | 7541/33185 (22.7) | 1.56 (1.49,1.64) |
| Any alcohol during pregnancy | 2071/36576 (5.7) | 297/3347 (8.8) | 1774/33229 (5.3) | 1.66 (1.48,1.87) |
| **Location of residence** |  |  |  |  |
| Top End, regional or remote | 23397/38347 (610) | 1848/3695 (50.0) | 21549/34652 (62.2) | ref |
| Top End, very remote | 7236/38347 (18.9) | 1084/3695 (29.3) | 6152/34652 (17.8) | 1.90 (1.75,2.05) |
| Central desert, remote | 4532/38347 (11.8) | 353/3695 (9.6) | 4179/34652 (12.1) | 0.99 (0.88,1.11) |
| Central desert, very remote | 3182/38347 (8.3) | 410/3695 (11.1) | 2772/34652 (8.0) | 1.63 (1.46,1.83) |
| **Pre-existing maternal risk factors, n(%)** |  |  |  |  |
| Anaemia | 1707 (44) | 240 (6.3) | 1467 (4.2) | 1.50 (1.32,1.71) |
| Diabetes | 673 (1.7) | 214 (5.6) | 459 (1.3) | 4.29 (3.66,5.03) |
| Hypertension | 349 (0.9) | 87 (2.3) | 262 (0.7) | 3.05 (2.40,3.88) |
| Cardiac disease | 924 (2.4) | 177 (4.6) | 747 (2.1) | 2.18 (1.85,2.56) |
| Renal disease | 406 (1.0) | 86 (2.2) | 320 (0.9) | 2.47 (1.95,3.13) |
| **Obstetric complications, n(%)** |  |  |  |  |
| Gestational anaemia | 2294 (5.9) | 249 (6.5) | 2045 (5.8) | 1.12 (0.99,1.27) |
| Gestational diabetes | 4175 (10.7) | 489 (12.7) | 3686 (10.4) | 1.22 (1.12,1.33) |
| Pre-eclampsia | 1316 (3.4) | 415 (10.9) | 901 (2.6) | 4.24 (3.79,4.74) |
| Antepartum haemorrhage | 714 (1.8) | 281 (7.3) | 433 (1.2) | 5.97 (5.16,6.91) |
| Intrauterine growth restriction | 1181 (3.0) | 348 (9.1) | 833 (2.4) | 3.84 (3.41,4.33) |
| Urinary tract infection | 1091 (2.8) | 160 (4.2) | 931 (2.6) | 1.58 (1.34,1.86) |
| Multiple pregnancy | 913 (2.3) | 567 (14.8) | 346 (1.0) | 15.07 (13.24,17.15) |
| **Birth characteristics:** |  |  |  |  |
| Born in the private hospital | 6619/38917 (17.0) | 455/3827 (11.9) | 6164/35090 (17.6) | 0.68 (0.62,0.74) |
| Induced birth | 9999 (25.5) | 658 (17.1) | 9341 (26.5) | 0.65 (0.60,0.70) |
| **Mode of delivery** |  |  |  |  |
| Vaginal | 26813 (68.5) | 2200 (57.3) | 24613 (69.7) | ref |
| Elective Caesar | 5816 (14.9) | 421 (11.0) | 5395 (15.3) | 0.88 (0.79,0.98) |
| Emergency Caesar | 6519 (16.7) | 1218 (32.7) | 5301 (15.0) | 2.28 (2.11,2.45) |
| **Infant characteristics** |  |  |  |  |
| First Nations baby | 14907 (38.1) | 2183 (56.9) | 12724 (36.0) | 1.58 (1.53,1.63) |
| Female | 19050 (48.7) | 1816 (47.3) | 17234 (48.8) | 0.97 (0.94,1.00) |
| Small for gestational age | 3900/39145 (10.0) | 552/3838 (14.4) | 3348/35307 (9.5) | 1.52 (1.40,1.65) |
| Admitted to the special care nursery | 6851/38860 (17.6) | 3020/3827 (78.9) | 3831/35033 (10.9) | 7.22 (7.00,7.47) |
| Died at age <1 month | 185 (0.5) | 152 (4.0) | 33 (0.1) | 42.36 (29.12,61.63) |
| Died at age <12 months | 250 (0.6) | 178 (4.6) | 72 (0.2) | 22.74 (17.33,29.84) |

**CI:** Confidence interval. **NT:** Northern Territory. Small for gestational age represents the proportion of infants in the lowest weight decile for gestational age (in weeks) calculated against national standards.

Compared to term infants, preterm infants were more likely to be First Nations, have mothers <19 years old, mothers who smoked or consumed alcohol, and mothers who resided in very remote areas. Preterm infants were less likely to be born at the private hospital or to mothers who received first-trimester antenatal care. Preterm birth was also associated with maternal diabetes, hypertension, renal disease, pre-eclampsia, intrauterine growth restriction, multiple pregnancy. Preterm infants had a higher infant mortality in the first 12 months (Table 2).

### Risk of RSV-ALRI hospitalisation

There were 881 RSV-ALRI hospitalisations among 846 infants (2.1% of the cohort, Table 3). The majority (812, 96.0%) experienced a single episode, while 33 (3.9%) infants had two episodes, and one infant (0.1%) had three. RSV-ALRI was the primary diagnosis in almost all cases (748/846, 88.4%). The median age at first admission was 130 days (IQR 68–209; Supplementary Table 1). For all cause ALRI, there were 5,192 episodes among 3,576 infants (9.1% of the cohort). Most infants (2614, 73.1%) had one episode, 608 (17.0%) had two, 201 (5.6%) had three, and 153 (4.3%) had more than three episodes. ALRI was the primary diagnosis in 91% of cases. The median age at first admission was 147.5 days (interquartile range [IQR] 61–213 days).

**Table 3.** Proportion of children hospitalised with RSV-ALRI and other respiratory conditions in the first year of life (2008-2017): by preterm birth status.

|  | Episodes | Proportion with condition<br>(N=39148) | Proportion with condition by preterm birth status |  | RR<br>(95% CI) | aRR1<br>(95% CI) | aRR2<br>(95% CI) |
| --- | --- | --- | --- | --- | --- | --- | --- |
|  |  |  | Yes<br>(N=3839) | No<br>(N=35309) |  |  |  |
|  | n | n (%) | n (%) | n (%) |  |  |  |
| RSV-ALRI, n (%) | 881 | 846 (2.1) | 167 (4.4) | 679 (1.9) | 2.26 (1.90,2.69) | 1.72 (1.43,2.08) | 1.83 (1.52,2.19) |
| ALRI, n (%) | 5192 | 3576 (9.1) | 644 (16.8) | 2932 (8.3) | 2.02 (1.86,2.19) | 1.44 (1.32,1.57) | 1.46 (1.34,1.49) |
| *Viral pneumonia* | 478 | 447 (1.1) | 80 (2.1) | 367 (1.0) | 2.00 (1.57,2.55) | 1.29 (0.98,1.69) | 1.36 (1.06,1.74) |
| *Bacterial pneumonia* | 1066 | 904 (2.3) | 163 (4.3) | 741 (2.1) | 2.02 (1.71,2.39) | 1.23 (1.03,1.48) | 1.25 (1.05,1.49) |
| *Bronchiolitis* | 3837 | 2798 (7.2) | 534 (13.9) | 2264 (6.4) | 2.17 (1.98,2.38) | 1.56 (1.41,1.72) | 1.61 (1.46,1.77) |
| *Bronchitis* | 27 | 20 (0.1) | 4 (0.1) | 16 (0.1) | 2.30 (0.77,6.87) | na | na |
| *Non-specific ALRI* | 515 | 479 (1.2) | 97 (2.5) | 382 (1.1) | 2.34 (1.87,2.92) | 1.65 (1.30,2.10) | 1.60 (1.27,2.00) |

Preterm-born infants had twice the risk of RSV-ALRI hospitalisation (4.4% vs. 1.9%; RR 2.26, 95% CI 1.90–2.69) compared to term-born infants (Table 3), with the highest risk observed in those born at 28–31 (7.6%) and 20-27 (5.5%) weeks gestation (Supplementary Table 2). The cumulative proportion of RSV-ALRI hospitalisations diverged between preterm and term infants from 1 to 12 months of age (Figure 2A). Frequency of first RSV-ALRI admissions peaked earlier in term infants (3 months) compared to preterm infants (5 months) (Figure 2B). Preterm birth was also associated with higher risks of hospitalisation for all-cause ALRI, viral and bacterial pneumonia, bronchiolitis, and non-specific ALRI (Table 3). Bronchitis diagnoses were rare (<1%) and excluded from further analysis.

**Figure 2.**
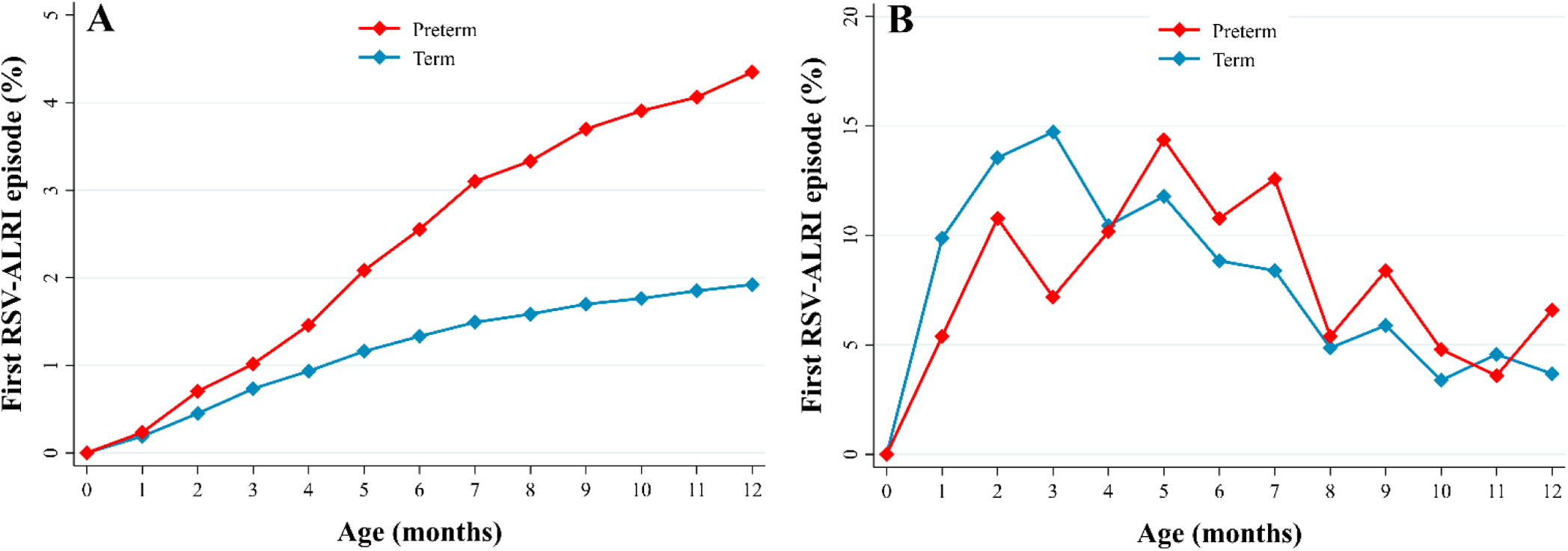
Monthly cumulative proportion (A) and frequency distribution (B) of RSV-ALRI hospitalisations among infants in the Northern Territory during their first year of life: Preterm (<37 weeks) and Term (≥37 weeks) born infants. **RSV-ALRI:** Respiratory syncytial virus associated acute lower respiratory infection. Frequency distribution is relative to total episodes (monthly episodes/total episodes)

Given that preterm birth and infant respiratory infections share overlapping risk factors, multivariable analyses were conducted to account for potential confounders that were available in the NT Midwives Collection (Table 3; Supplementary Table 3). After adjusting for general maternal characteristics, pre-existing health risks, obstetric complications, and birth/infant characteristics, the positive association between preterm birth and RSV-ALRI hospitalisations remained statistically significant, though attenuated (aRR1 1.73, 95% CI 1.43–2.08; aRR2 1.83, 95% CI 1.52–2.19). Similarly, significant associations persisted for all-cause ALRI, bronchiolitis, and non-specific ALRI after adjustment, but the associations with viral and bacterial pneumonia were either not significant or weak.

The full multivariable model (Supplementary Table 3) identified several additional factors associated with a higher risk of RSV-ALRI hospitalisation. These included being a First Nations infant (aRR 2.40, 95% CI 1.97–2.93), having a mother younger than 19 years (aRR 1.80, 95% CI 1.40–2.30, compared to mothers aged 30–39), living in the very remote Top End (aRR 1.61, 95% CI 1.31–1.98), or in remote (aRR 2.72, 95% CI 2.18–3.38) or very remote (aRR 3.61, 95% CI 2.93–4.30) Central Desert regions (compared to Top End regional or remote areas), and having a mother with a urinary tract infection during pregnancy (aRR 1.45, 95% CI 1.10–1.89). Conversely, female infants (aRR 0.79, 95% CI 0.69–0.91), infants of primiparous women (aRR 0.65, 95% CI 0.55–0.78), and infants whose mothers received antenatal care in the first trimester (aRR 0.86, 95% CI 0.74–1.01) had a lower risk of RSV-ALRI hospitalisation. Univariate comparisons of RSV-ALRI hospitalisations by these characteristics are presented for reference in Supplementary Tables 4A, 4B, 4C. Notably, the risk factors were largely consistent across all respiratory conditions. However, preeclampsia was associated with increased risks of all-cause ALRI, pneumonia, and bronchiolitis, while first-trimester antenatal care reduced these risks. Maternal smoking and alcohol use were weakly associated with all-cause ALRI and bronchiolitis.

The pseudo R² values from the multivariable models (Supplementary Table 3) indicate that, while the models capture important associations, a substantial proportion of the variation in outcomes remains unexplained.

Finally, to determine whether preterm birth was a specific risk factor for RSV-ALRI or a general risk factor for poor health, we also evaluated its association with several other hospitalised conditions (Supplementary Tables 5 and 6). In the multivariable analyses, preterm birth showed minimal associations with gastroenteritis (aRR1 1.12, 95% CI 0.99–1.27; aRR2 1.16, 95% CI 1.03–1.31), skin infection (aRR1 1.04, 95% CI 0.87–1.25; aRR2 1.09, 95% CI 0.92–1.29), and scabies (aRR1 1.19, 95% CI 1.00–1.42; aRR2 1.16, 95% CI 0.98–1.36).

Notably, infants living in the very remote Central Desert region (compared to those in regional or remote Top End areas) had the highest risk of hospitalisation for gastroenteritis (aRR 4.44, 95% CI 3.92–5.02), skin infection (aRR 2.88, 95% CI 2.39–3.46), and scabies (aRR 5.71, 95% CI 4.45–7.32). Additionally, First Nations infants had a markedly higher risk of hospitalisation with a diagnosis of scabies (aRR 32.17, 95% CI 19.76–52.36) compared to other children.

### Temporal trends in RSV-ALRI hospitalisations

After excluding data from 2008 to avoid lead-in truncation bias, the average annual incidence of RSV-ALRI hospitalisations over the decade was 2.3 cases per 100 child-years (Figure 3 and Supplementary Table 7). The rate decreased by about 9% each year (incidence rate ratio [IRR] 0.91 per year, 95% CI 0.86–0.95). Similar downward trends were observed for both viral and bacterial pneumonia. In contrast, the incidence rates of bronchiolitis, non-specific ALRI, and all cause ALRI remained relatively stable.

**Figure 3.**
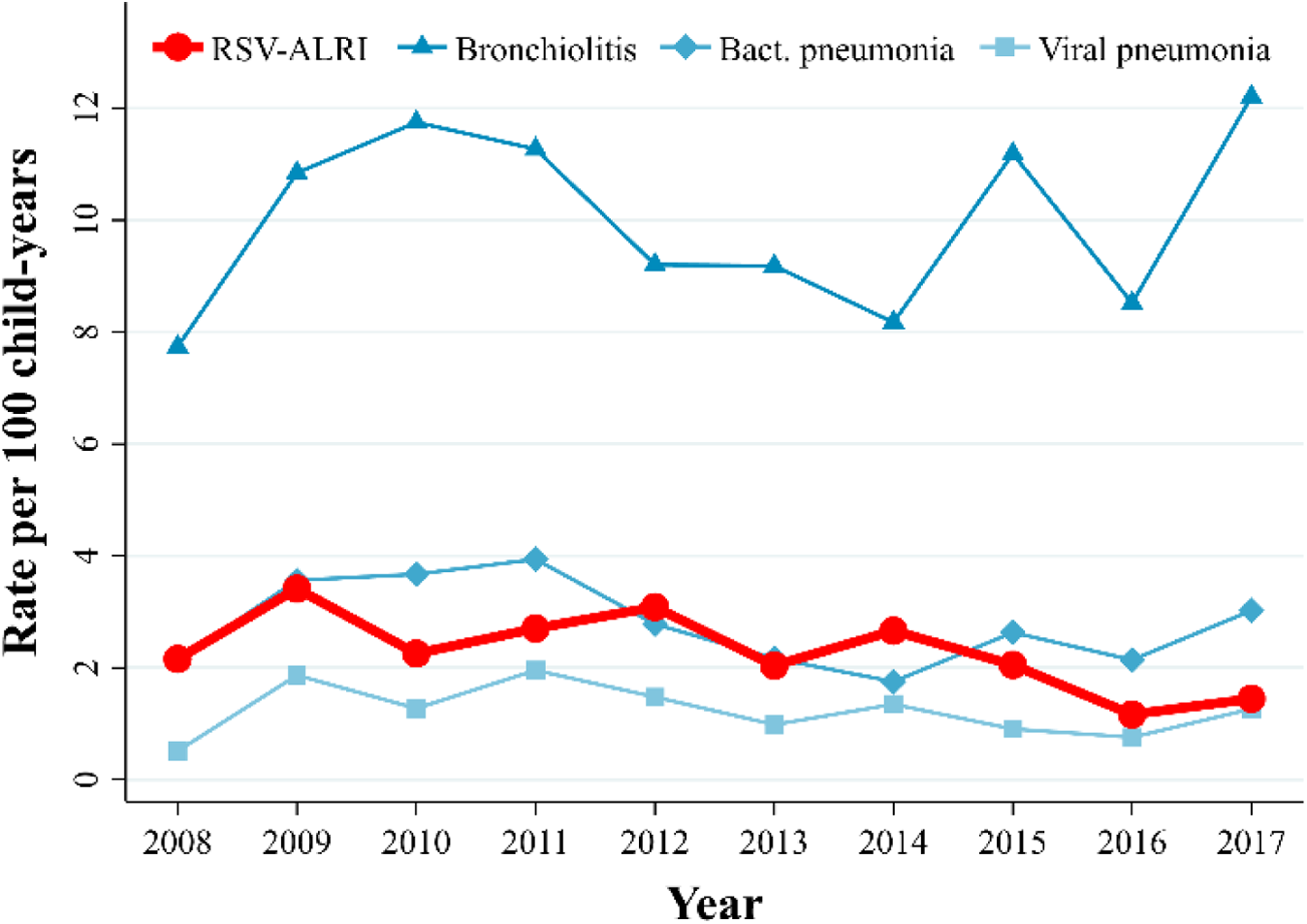
Annual hospitalisation rates for RSV-ALRI and other respiratory infection classifications among Northern Territory infants: 2008-2017. **RSV-ALRI:** Respiratory syncytial virus associated acute lower respiratory infection.

Adjusting for year of hospitalisation episode, the average annual incidence rate of RSV-ALRI hospitalisation was higher among preterm compared to term infants (IRR 2.33, 95% CI 1.87– 2.92), higher among infants residing in the Central Desert compared to those in the Top End (IRR 3.43, 95% CI 2.76–4.27), and higher among First Nations compared to non-First Nations infants (IRR 4.59, 95% CI 3.48-6.06). There was no significant interaction between time and preterm birth status, nor between time and residing in the Central Desert region, indicating that the time trends in RSV-ALRI were similar for these groups. However, the interaction between being a First Nations infant and time approached significance (interaction IRR 0.92, 95% CI 0.84–1.01), suggesting a closing of the gap in RSV-ALRI rates between First Nations and non-First Nations infants over time. In models including this interaction, the main effect of time was no longer significant supporting the validity of this finding.

Interestingly, the frequency of first RSV-ALRI admissions among non-First Nations children was highest in the first three months of life, whereas for First Nations infants, admissions were elevated and sustained across the first eight months of life (Supplementary Figure 3). The frequency of admissions by infant month of age did not differ between Central Desert and Top End regions (Supplementary Figure 4). However, there were distinct seasonal trends by region: in the Top End, aggregate RSV-ALRI cases per month (summed over the decade) peaked from December to April, while in the Central Desert, cases were highest from June to October (Supplementary Figure 5).

## Discussion

This is the first study to evaluate the association between preterm birth and the risk of infant RSV-ALRI hospitalisations in Australia’s NT. Aligning with our hypothesis, preterm birth was associated with an increased risk of RSV-ALRI hospitalisation in the first year of life. Furthermore, this work highlights the disproportionate burden of RSV-ALRI among First Nations infants and those residing in remote regions, emphasizing an intersection between biological vulnerability (preterm) and social determinants of health.

The period prevalence of preterm birth (10%) demonstrated in this NT-wide study was consistent with earlier studies in the Top End (10%)^1,13^ and national trends (9%).^14^ First Nations families had a 2.1-times higher risk of preterm birth than non-First Nations families at a rate (14.6%) well above the global average (11.1%).^15^ The key factors linked to preterm birth in our study included preexisting diabetes and hypertension, pre-eclampsia, antepartum haemorrhage, intrauterine growth restriction, multiple pregnancy. Apart from multiple pregnancy, these risk factors have been identified as more common among First Nations women. ^1^

Infants born preterm had 1.7-1.8 times the risk of RSV-ALRI hospitalisation than term-born infants after adjusting for demographic and antenatal health factors. The risk was most pronounced for infants born <31 weeks gestation, aligning with global trends. A multinational meta-analysis spanning 52 countries (including 13 low- and middle-income nations) found that preterm infants account for 25% of all RSV hospitalisations. ^2^ The biological link between preterm birth and severe respiratory infections is driven by underdeveloped airways and immune function, reduced maternal antibody transfer, chronic lung conditions such as bronchopulmonary dysplasia, microbiome dysbiosis, increased oxidative stress, and genetic or epigenetic factors, all of which heighten susceptibility. ^16^

Preterm birth is an important factor when planning RSV prevention. From February 2025, all pregnant women in Australia are eligible for a free dose of maternal RSV vaccine (Pfizer) between 28 and 36 weeks gestation, ideally administered at least two weeks before delivery to optimise antibody transfer. Earlier vaccination may benefit those at risk of preterm birth. While some studies suggested a possible link between maternal RSV vaccination and preterm birth, especially in lower-middle income settings, ^17–19^ prelicensure^20^ and early post-marketing studies^21^ have not shown an increased risk when the vaccine is given from 32 weeks onwards. If birth occurs before vaccination, or within two weeks of vaccination, long-acting monoclonal antibody (nirsevimab) is recommended as per national guidelines to ensure infant protection.^22^

Preterm birth was one of several risk factors for early-onset RSV-ALRI hospitalisation in the NT. Other prominent predictors included younger maternal age, residence in remote and Central Desert regions, maternal urinary tract infection (UTI) in pregnancy and male sex. While younger maternal age and male sex align with prior studies, ^6^ this is the first report suggesting maternal UTI to increased RSV-ALRI hospitalisation risk in infants. Notably, our multivariable analysis showed that the UTI association was independent of preterm birth status, suggesting distinct biological pathways. While further exploration of this potential association is needed, urinary and genital tract infections have been shown to complicate pregnancy through inflammatory pathways and disruption of the vaginal microbiome, ^23^ which shapes the neonatal gut and lung microbiome^24,25^ – critical for immune development and resilience against infections. ^26^ Interestingly, maternal smoking was not a risk factor for infant RSV hospitalisation after accounting for preterm birth.

RSV-ALRI hospitalisation rates in the NT declined between 2008-2017, with a more marked reduction among First Nations infants suggesting some progress in narrowing the health gap. However, changes in testing, coding, health-care initiatives, and random variation may have influenced these trends, and ongoing surveillance in the post-COVID era is needed for confirmation. Historically, First Nations communities in Australia and internationally have experienced a disproportionate burden of RSV-ALRI. ^3,7,27–29^ Across the decade, First Nations infants had 2.3-times the (adjusted) per child risk of RSV-ALRI hospitalisation than others. This disparity reflects complex factors, including limited access to culturally safe health care, socioeconomic disadvantage, and systemic racism. Continued efforts are essential to achieve health equity for all children.

Our study confirms a regional contrast in RSV seasonality across the NT. In the Top End, RSV-ALRI hospitalisations peaked during the wet season, consistent with earlier studies showing correlations with rainfall and humidity. ^4^ In the Central Desert, peaks occurred in the cooler winter and spring months, despite low rainfall and humidity, a pattern more typical of temperate southern Australia. This paradox likely reflects different dominant drivers. In the tropics, humidity promotes viral stability^30^ while in arid and temperate zones, lower temperatures and lower humidity enhance viral survival and transmission. ^31^ In both regions, indoor crowding is more common during these periods. These findings indicate that both environmental and social factors shape RSV seasonality across diverse climates.

Strengths of this study include the large, population-based cohort and robust data linkage, enabling comprehensive analysis across diverse contexts. Limitations include reliance on ICD-10AM codes, which, while highly specific for RSV, may underestimate true burden due to low sensitivity (∼25%) and diagnostic imprecision, especially for single admissions or non-specific codes. ^32,33^

## Conclusion

Early-life RSV-ALRI infection can have immediate (hospitalisation and death) and long-term consequences (wheezing, asthma and impaired lung function into adulthood). As such, prevention of infant RSV infections is a global priority. This study demonstrates that preterm birth is a significant risk factor for RSV-ALRI hospitalisation among infants in the NT. First Nations families and those living in remote-areas bear a disproportionate burden of both preterm birth and RSV-ALRI. Addressing these disparities is complex and requires a multifaceted approach, including First Nations designed antenatal care and child health programs alongside tailored RSV prophylaxis and management for preterm born infants. Over the last 15 years culturally enhanced midwifery group practice, ^34^ smoking cessation, ^35^ cervical length screening^36^ and birthing on country programs^37,38^ have been introduced to tackle preterm birth among NT First Nations families and rates have remained stable over this time. The recent national roll out of maternal RSV vaccines and infant immunisation therapies is an important advance in RSV prevention. ^22^ Monitoring uptake and effectiveness will be crucial to ensure adequate protection to all preterm born and at-risk infants.

## Data Availability

All summary statistics are available on request. Deidentified individual participant data will not be made available.

## Ethics

The project was approved by the NT Human Research Ethics Committee (2019-3551, 2018-3261) and Aboriginal Ethics Subcommittee. Individual-level consent was waived for this deidentified data. The project was also approved by First Nations Advisory Groups from Menzies School of Health Research and Child and Youth Development Research Partnership.

## Acknowledgments

This project was supported by the Child and Youth Development Research Partnership (CYDRP), a collaboration between the Menzies School of Health Research and the Northern Territory Government that uses linked data to study child and adolescent outcomes. We thank SA-NT DataLink for coordinating the data linkage, and the Northern Territory Department of Health for providing access to their data through CYDRP. We also acknowledge and thank all the NT Aboriginal and Torres Strait Islander children and their families involved in the study. The views expressed in this article are those of the authors and do not necessarily reflect the views of the data custodians.

---

**Supplementary Figure 1.**
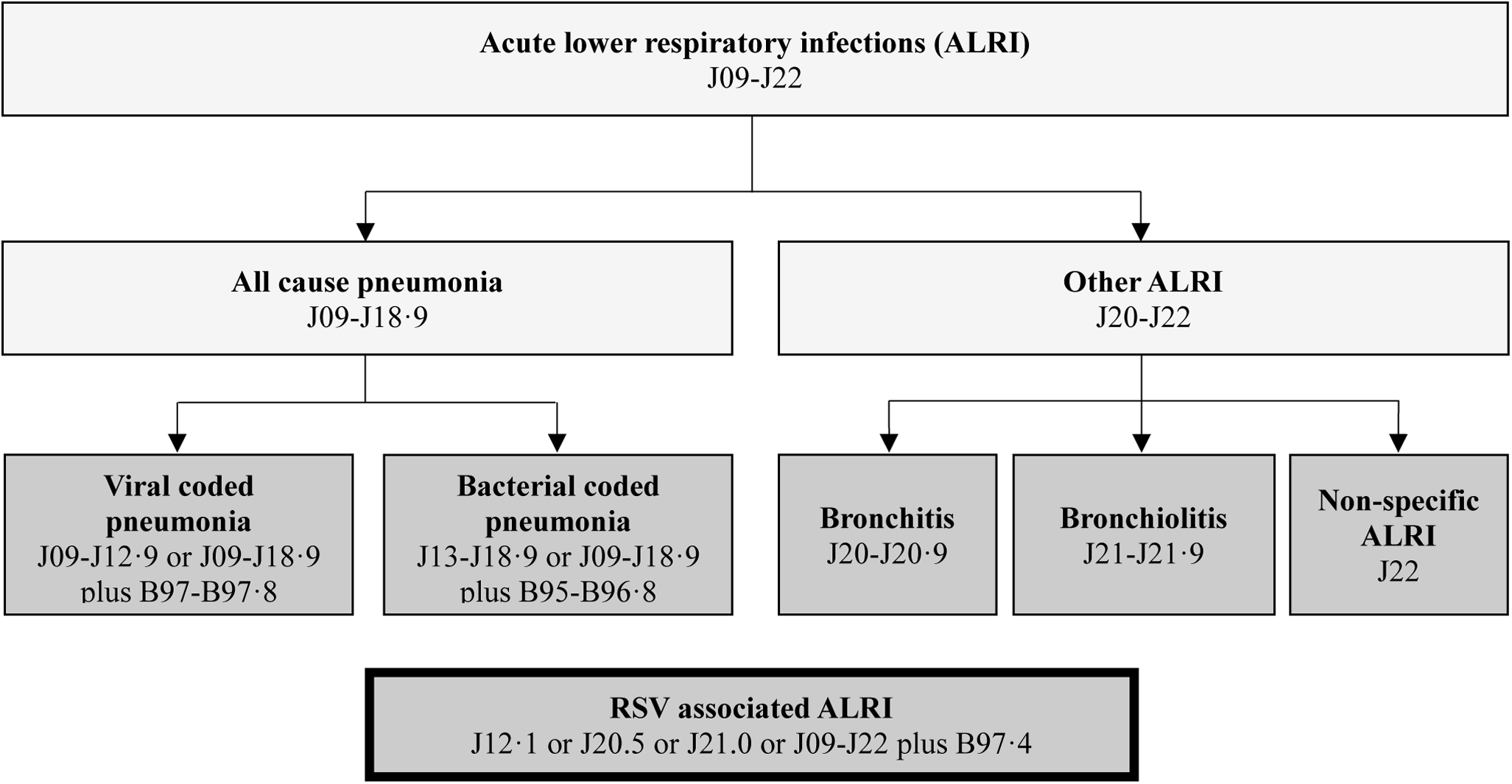
ICD10 diagnostic categories. International classification of diseases codes (tenth revision, Australian modification; ICD-10AM) were used to identify Respiratory syncytial virus (RSV)-associated ARI (the primary outcome) and other respiratory hospital outcomes of interest. The study data were sourced from the Hospital Trends Dataset. This dataset is derived from the Northern Territory Inpatient Activity collection but is truncated to contain a maximum of ten ICD codes per hospital admission episode. All codes within an episode contributed to the diagnosis for that episode. As such, diagnoses were not always mutually exclusive within an episode. General respiratory classifications (J09-J22) and subgroups were supplemented by specific pathogen/disease codes; B95-B96·8: All bacterial codes as the cause of diseases in other chapters; B97-B97·8: All viral codes as the cause of diseases in other chapters; B97·4: RSV as the cause of diseases in other chapters.

**Supplementary Figure 2.**
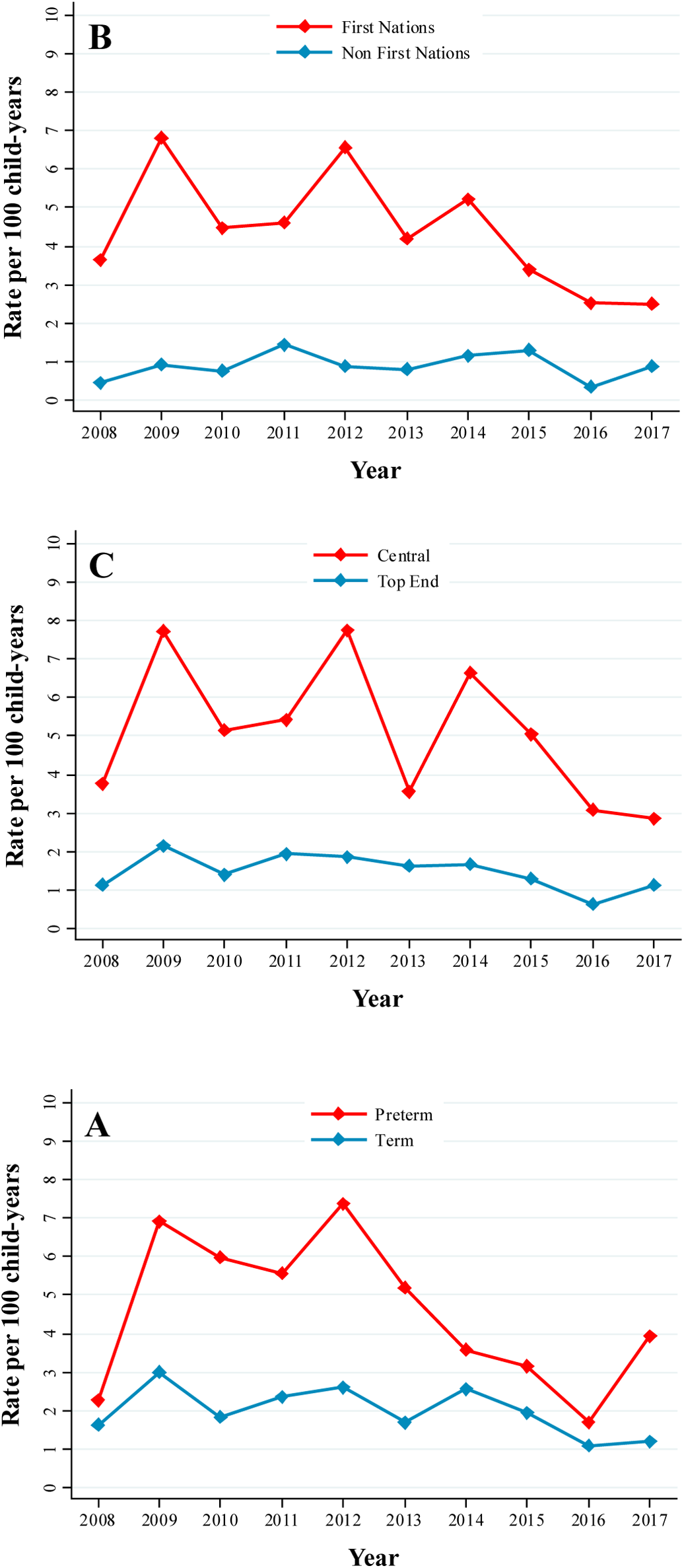
Annual RSV-ALRI hospitalisation rates in the first year of life: preterm and term. **(A)**, **First Nations and non-First Nations infants (B), Top End and Central desert (C)**

**Supplementary Figure 3.**
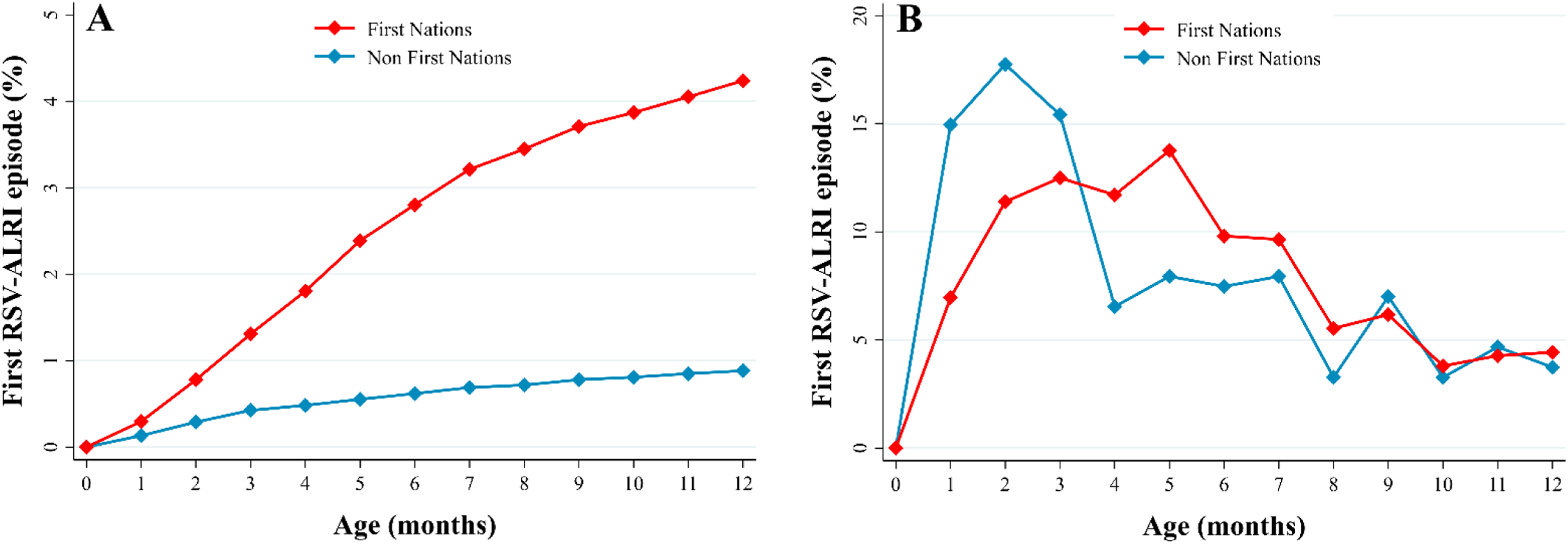
Monthly cumulative proportion (A) and frequency distribution (B) of the first RSV-ALRI hospitalisations in the first year of life: First Nations and non-First Nations infants.

**Supplementary Figure 4.**
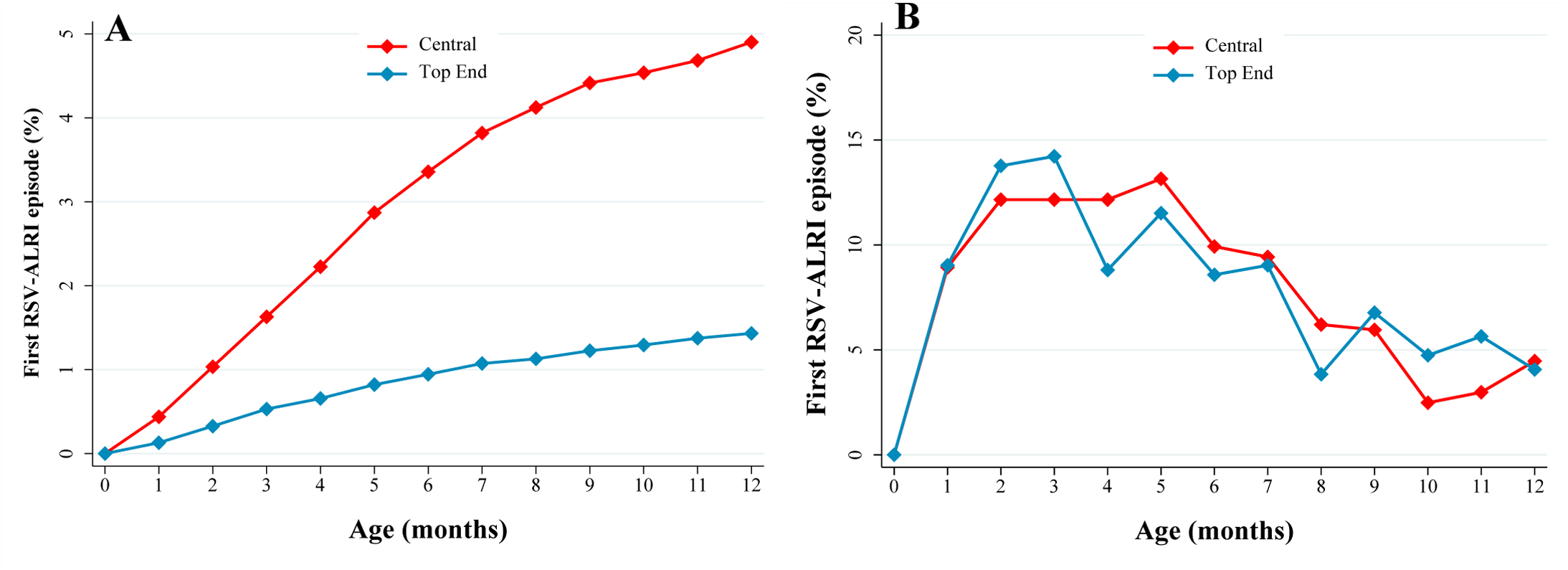
Monthly cumulative proportion (A) and frequency distribution (B) of the first RSV-ALRI hospitalisations in the first year of life: Top End and Central desert regions.

**Supplementary Figure 5.**
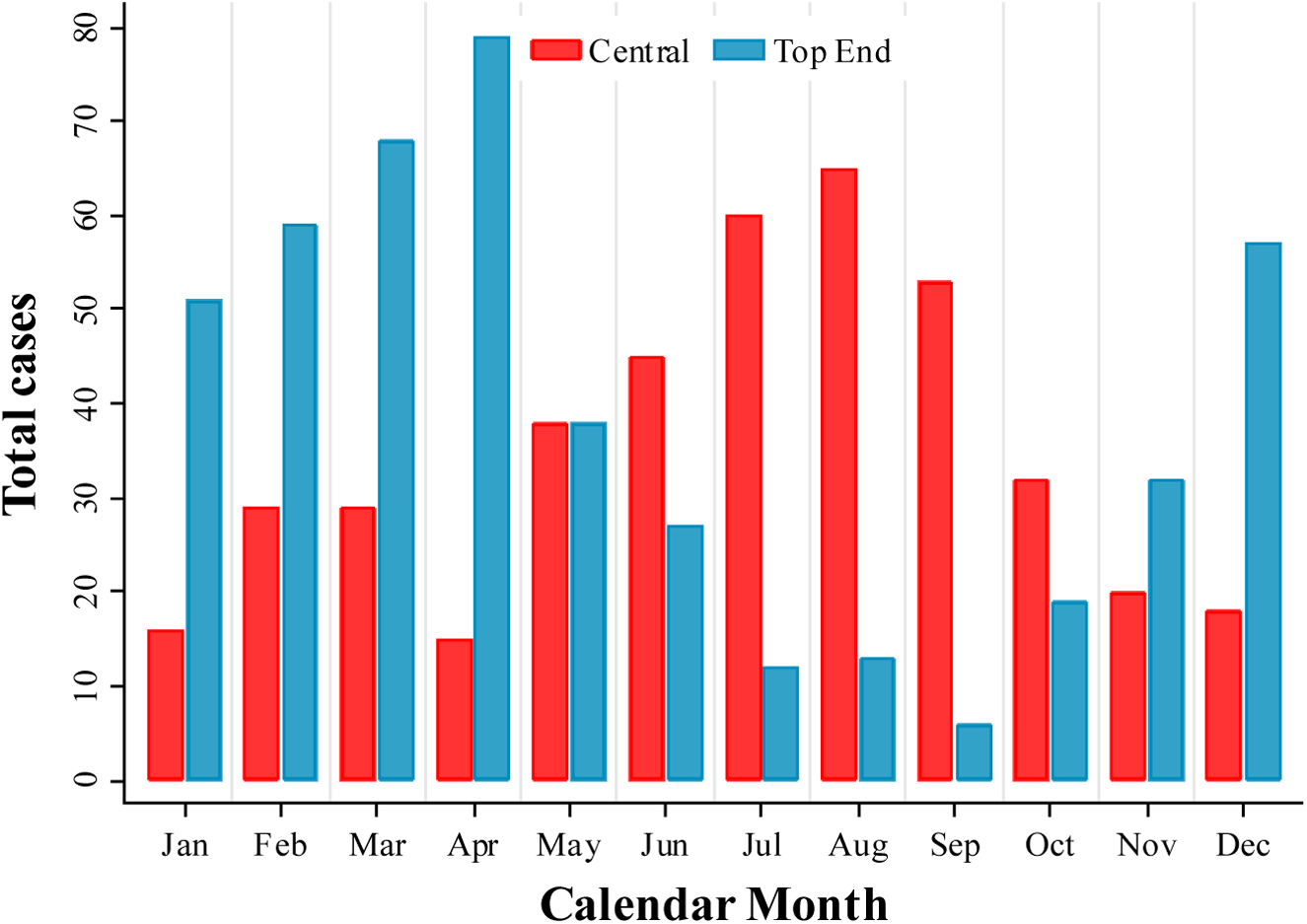
Total RSV-ALRI hospitalisations in the Northern Territory by calendar month over the decade 2008-2017: Top End and Central desert regions.

**Supplementary Table 1.**
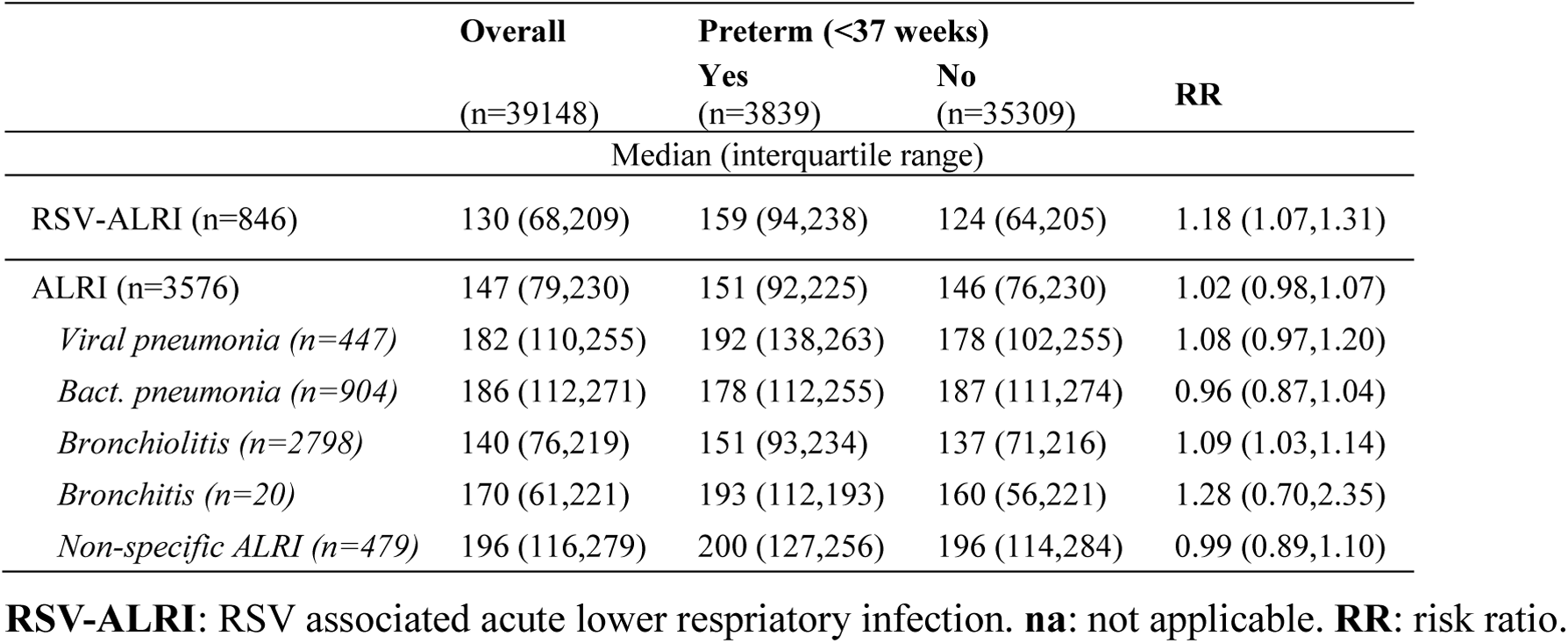
Age (in days) of first respiratory hospitalisations among Northern Territory infants in the first year of life: by preterm birth status.

|  | Overall<br>(n=39148) | Preterm (<37 weeks) |  | RR |
| --- | --- | --- | --- | --- |
|  |  | Yes<br>(n=3839) | No<br>(n=35309) |  |
|  | Median (interquartile range) |  |  |  |
| RSV-ALRI (n=846) | 130 (68,209) | 159 (94,238) | 124 (64,205) | 1.18 (1.07,1.31) |
| ALRI (n=3576) | 147 (79,230) | 151 (92,225) | 146 (76,230) | 1.02 (0.98,1.07) |
| *Viral pneumonia (n=447)* | 182 (110,255) | 192 (138,263) | 178 (102,255) | 1.08 (0.97,1.20) |
| *Bact. pneumonia (n=904)* | 186 (112,271) | 178 (112,255) | 187 (111,274) | 0.96 (0.87,1.04) |
| *Bronchiolitis (n=2798)* | 140 (76,219) | 151 (93,234) | 137 (71,216) | 1.09 (1.03,1.14) |
| *Bronchitis (n=20)* | 170 (61,221) | 193 (112,193) | 160 (56,221) | 1.28 (0.70,2.35) |
| *Non-specific ALRI (n=479)* | 196 (116,279) | 200 (127,256) | 196 (114,284) | 0.99 (0.89,1.10) |

**RSV-ALRI:** RSV associated acute lower respiratory infection. **na:** not applicable. **RR:** risk ratio.

**Supplementary Table 2.**
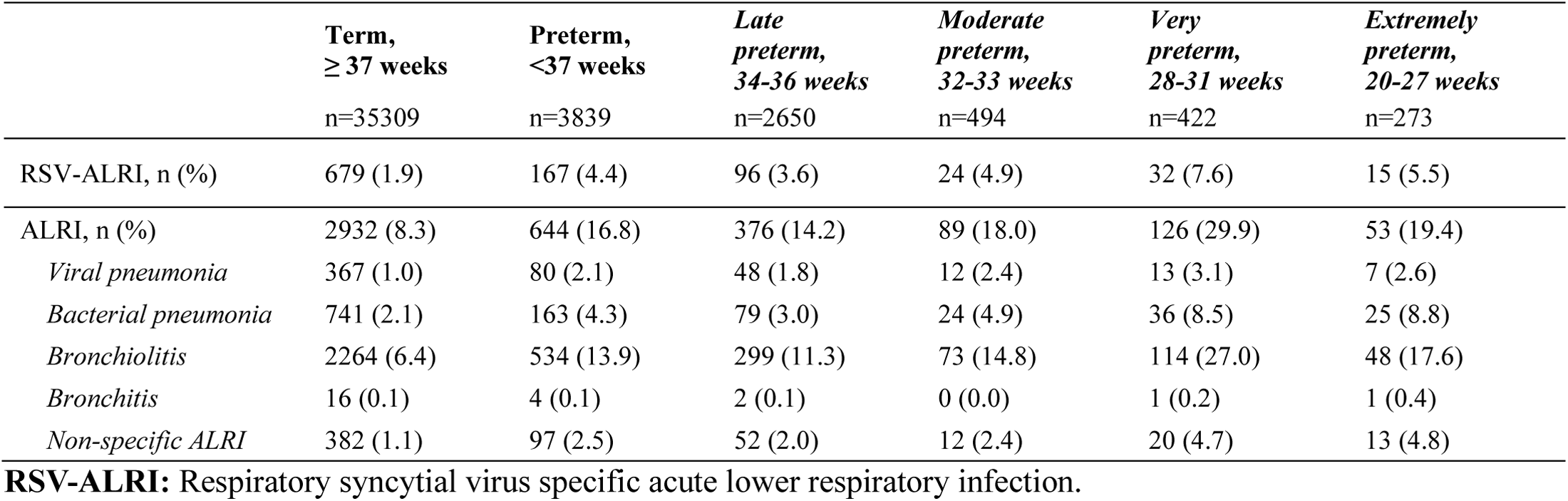
Prevalence of respiratory hospitalisations among Northern Territory infants in the first year of life: by preterm birth category.

**Supplementary Table 3.**
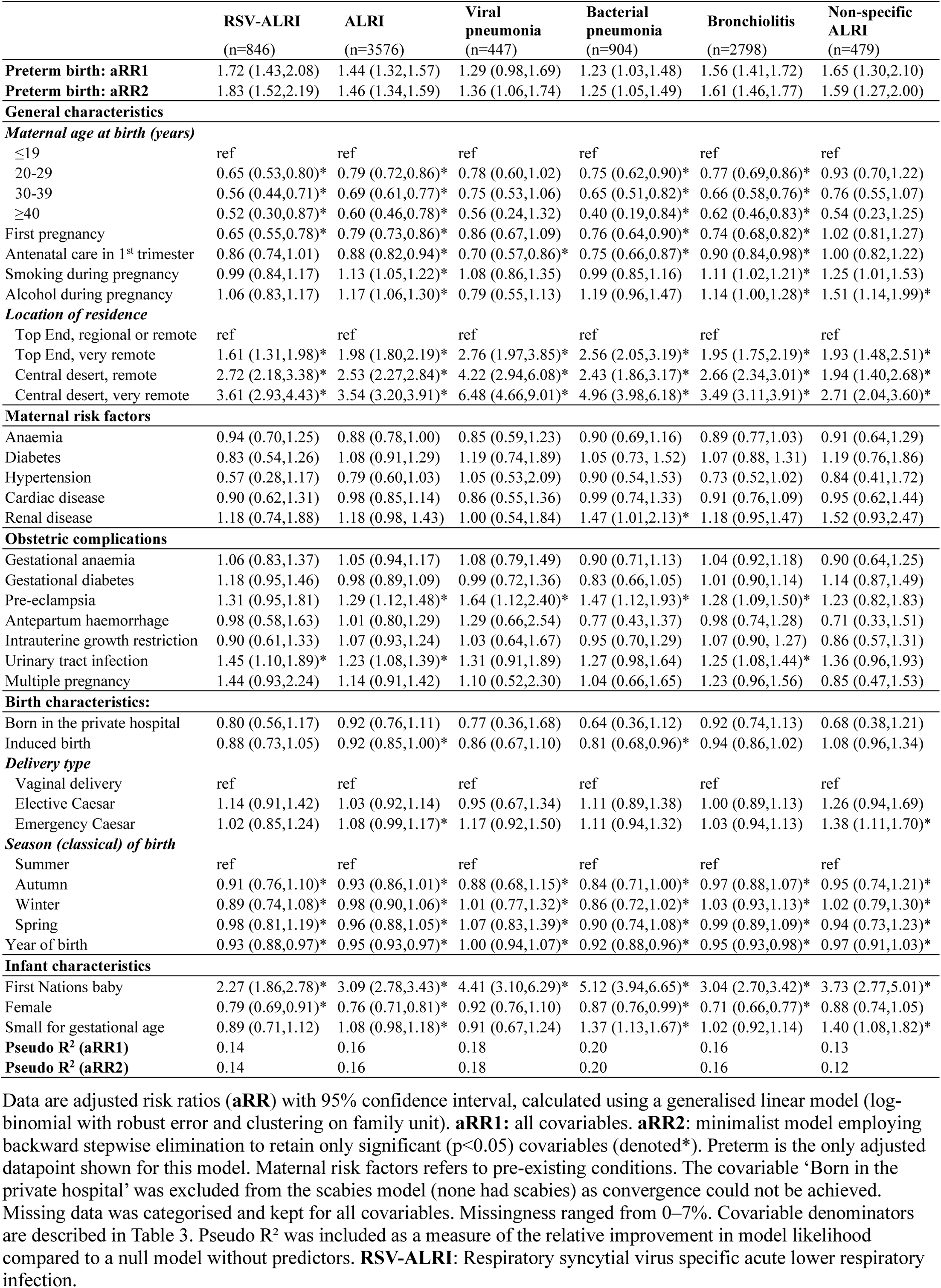
Multivariable model for preterm birth and infant respiratory hospitalisations.

**Supplementary Table 4.**
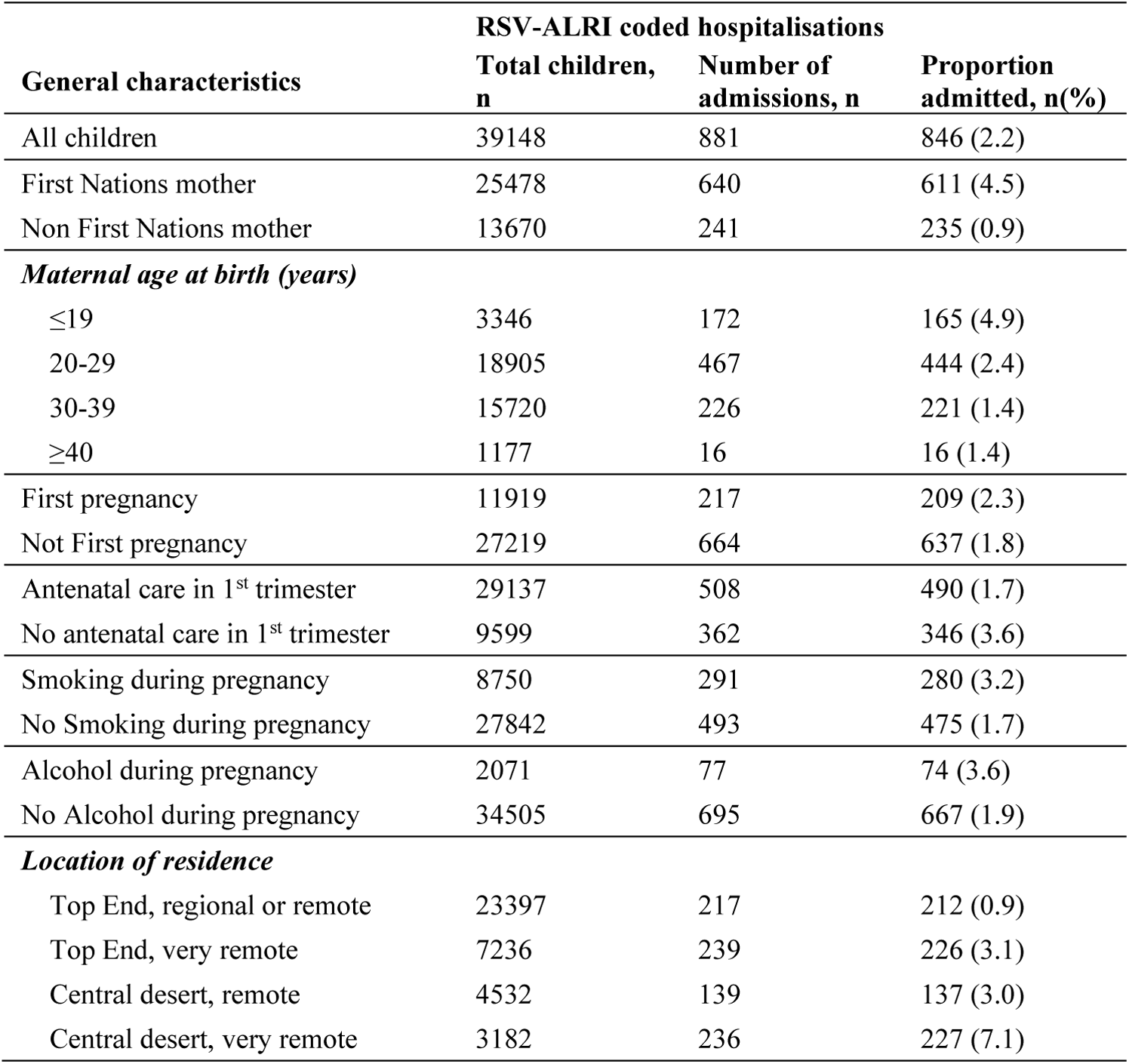
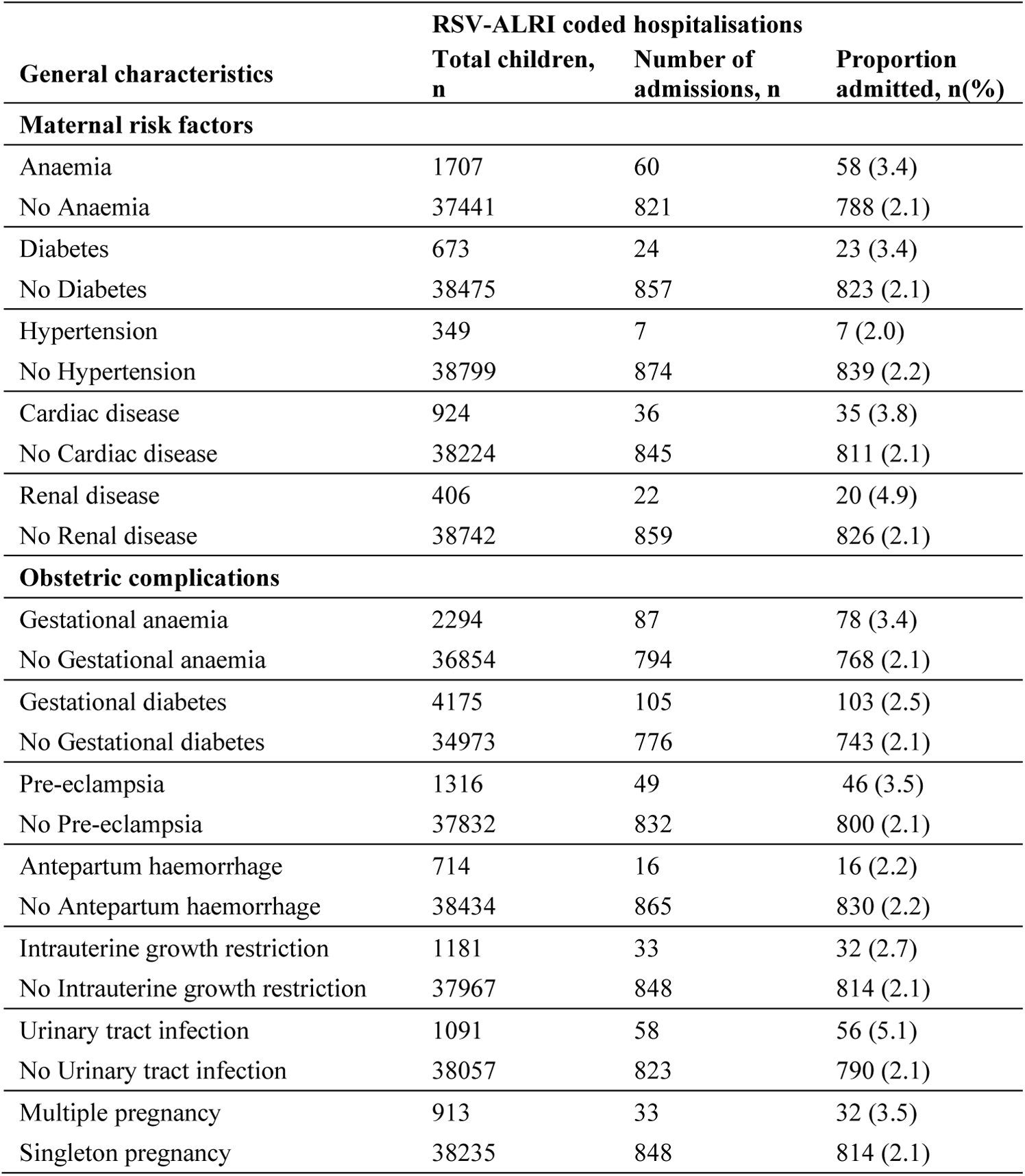
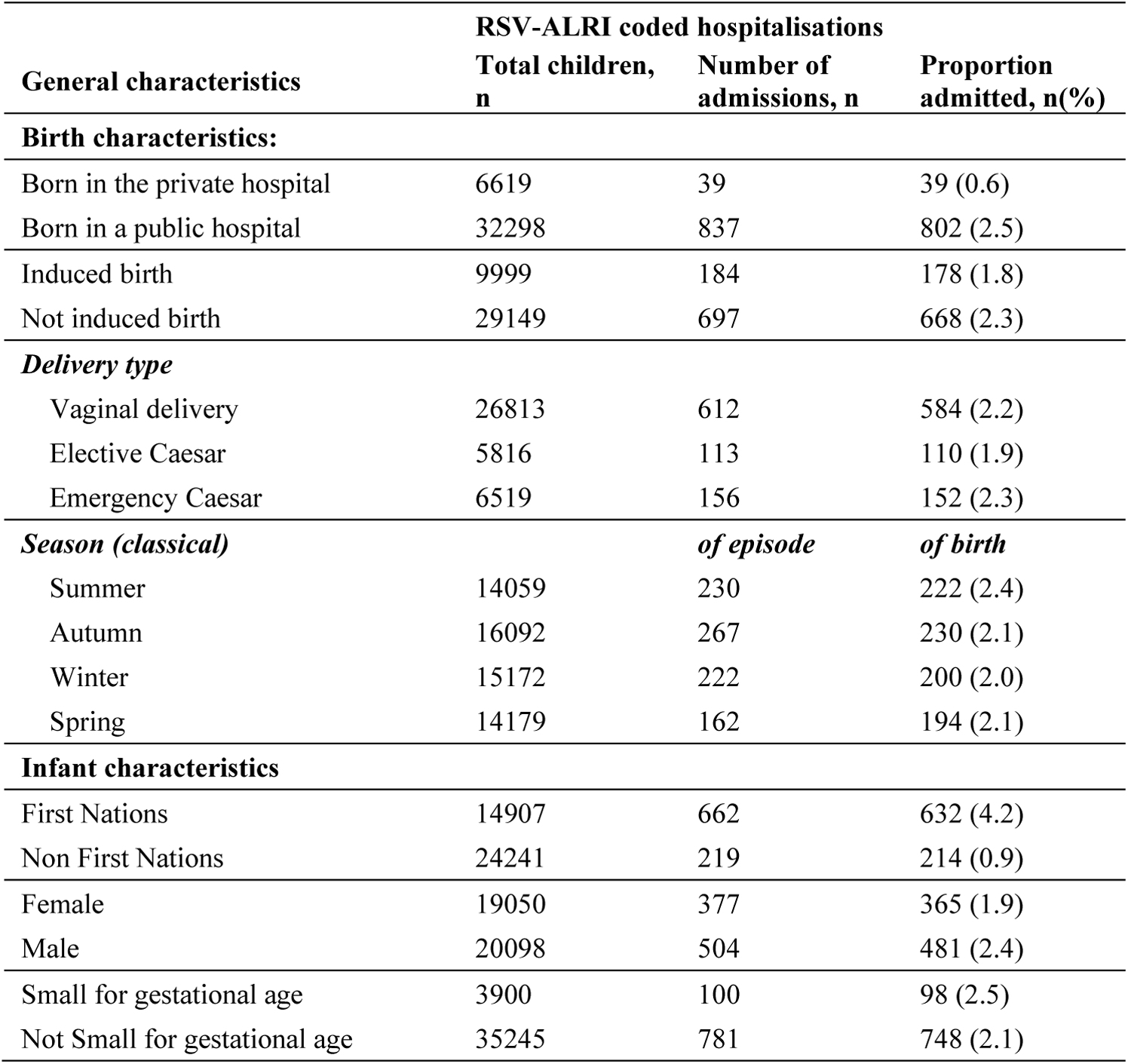
A. RSV-ALRI hospitlisations among Northern Territory infants in the first year of life: by key maternal demographic factors, B. RSV-ALRI hospitlisations among Northern Territory infants in the first year of life: by maternal (pre-existing) and obstetric risk factors, C. RSV-ALRI hospitlisations among Northern Territory infants in the first year of life: by birth and infant characteristics.

**Supplementary Table 5.** Prevalence of ‘other’ ICD10 coded conditions in the first year of life (2008-2017): by preterm birth status.

|  | Overall | Preterm<br>(<37 weeks) |  | RR | aRR1 | aRR2 |
| --- | --- | --- | --- | --- | --- | --- |
|  | (n=39418) | Yes<br>(n=3839) | No<br>(n=35309) | (95% CI) | (95% CI) | (95% CI) |
| Gastroenteritis | 2236 (5.7) | 331 (8.6) | 1905 (5.4) | 1.60 (1.42,1.80) | 1.12 (0.99,1.27) | 1.16 (1.03,1.31) |
| Skin infection | 1105 (2.8) | 160 (4.2) | 945 (2.7) | 1.56 (1.31,1.85) | 1.04 (0.87,1.25) | 1.09 (0.92,1.29) |
| Scabies | 1100 (2.8) | 197 (5.1) | 903 (2.6) | 2.01 (1.71,2.35) | 1.19 (1.00,1.42) | 1.16 (0.98,1.36) |

**Supplementary Table 6.**
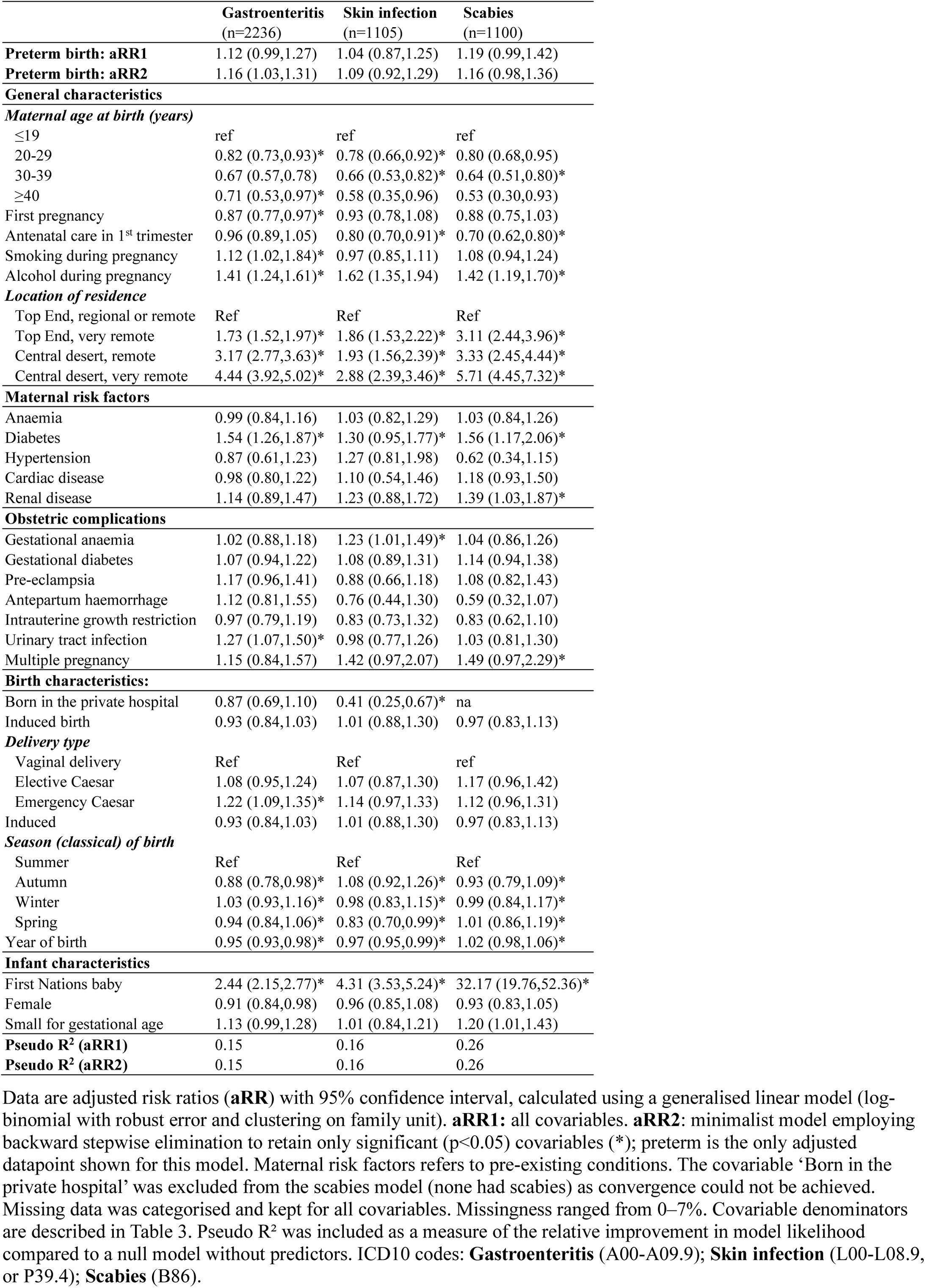
Multivariable model for preterm birth and ‘other’ infant hospitalisation outcomes.

|  | **Gastroenteritis**<br>(n=2236) | **Skin infection**<br>(n=1105) | **Scabies**<br>(n=1100) |
| --- | --- | --- | --- |
| **Preterm birth: aRR1** | 1.12 (0.99,1.27) | 1.04 (0.87,1.25) | 1.19 (0.99,1.42) |
| **Preterm birth: aRR2** | 1.16 (1.03,1.31) | 1.09 (0.92,1.29) | 1.16 (0.98,1.36) |
| **General characteristics** |  |  |  |
| *Maternal age at birth (years)* |  |  |  |
| ≤19 | ref | ref | ref |
| 20-29 | 0.82 (0.73,0.93)* | 0.78 (0.66,0.92)* | 0.80 (0.68,0.95) |
| 30-39 | 0.67 (0.57,0.78) | 0.66 (0.53,0.82)* | 0.64 (0.51,0.80)* |
| ≥40 | 0.71 (0.53,0.97)* | 0.58 (0.35,0.96) | 0.53 (0.30,0.93) |
| First pregnancy | 0.87 (0.77,0.97)* | 0.93 (0.78,1.08) | 0.88 (0.75,1.03) |
| Antenatal care in 1<sup>st</sup> trimester | 0.96 (0.89,1.05) | 0.80 (0.70,0.91)* | 0.70 (0.62,0.80)* |
| Smoking during pregnancy | 1.12 (1.02,1.84)* | 0.97 (0.85,1.11) | 1.08 (0.94,1.24) |
| Alcohol during pregnancy | 1.41 (1.24,1.61)* | 1.62 (1.35,1.94) | 1.42 (1.19,1.70)* |
| *Location of residence* |  |  |  |
| Top End, regional or remote | Ref | Ref | Ref |
| Top End, very remote | 1.73 (1.52,1.97)* | 1.86 (1.53,2.22)* | 3.11 (2.44,3.96)* |
| Central desert, remote | 3.17 (2.77,3.63)* | 1.93 (1.56,2.39)* | 3.33 (2.45,4.44)* |
| Central desert, very remote | 4.44 (3.92,5.02)* | 2.88 (2.39,3.46)* | 5.71 (4.45,7.32)* |
| **Maternal risk factors** |  |  |  |
| Anaemia | 0.99 (0.84,1.16) | 1.03 (0.82,1.29) | 1.03 (0.84,1.26) |
| Diabetes | 1.54 (1.26,1.87)* | 1.30 (0.95,1.77)* | 1.56 (1.17,2.06)* |
| Hypertension | 0.87 (0.61,1.23) | 1.27 (0.81,1.98) | 0.62 (0.34,1.15) |
| Cardiac disease | 0.98 (0.80,1.22) | 1.10 (0.54,1.46) | 1.18 (0.93,1.50) |
| Renal disease | 1.14 (0.89,1.47) | 1.23 (0.88,1.72) | 1.39 (1.03,1.87)* |
| **Obstetric complications** |  |  |  |
| Gestational anaemia | 1.02 (0.88,1.18) | 1.23 (1.01,1.49)* | 1.04 (0.86,1.26) |
| Gestational diabetes | 1.07 (0.94,1.22) | 1.08 (0.89,1.31) | 1.14 (0.94,1.38) |
| Pre-eclampsia | 1.17 (0.96,1.41) | 0.88 (0.66,1.18) | 1.08 (0.82,1.43) |
| Antepartum haemorrhage | 1.12 (0.81,1.55) | 0.76 (0.44,1.30) | 0.59 (0.32,1.07) |
| Intrauterine growth restriction | 0.97 (0.79,1.19) | 0.83 (0.73,1.32) | 0.83 (0.62,1.10) |
| Urinary tract infection | 1.27 (1.07,1.50)* | 0.98 (0.77,1.26) | 1.03 (0.81,1.30) |
| Multiple pregnancy | 1.15 (0.84,1.57) | 1.42 (0.97,2.07) | 1.49 (0.97,2.29)* |
| **Birth characteristics:** |  |  |  |
| Born in the private hospital | 0.87 (0.69,1.10) | 0.41 (0.25,0.67)* | na |
| Induced birth | 0.93 (0.84,1.03) | 1.01 (0.88,1.30) | 0.97 (0.83,1.13) |
| *Delivery type* |  |  |  |
| Vaginal delivery | Ref | Ref | ref |
| Elective Caesar | 1.08 (0.95,1.24) | 1.07 (0.87,1.30) | 1.17 (0.96,1.42) |
| Emergency Caesar | 1.22 (1.09,1.35)* | 1.14 (0.97,1.33) | 1.12 (0.96,1.31) |
| Induced | 0.93 (0.84,1.03) | 1.01 (0.88,1.30) | 0.97 (0.83,1.13) |
| *Season (classical) of birth* |  |  |  |
| Summer | Ref | Ref | Ref |
| Autumn | 0.88 (0.78,0.98)* | 1.08 (0.92,1.26)* | 0.93 (0.79,1.09)* |
| Winter | 1.03 (0.93,1.16)* | 0.98 (0.83,1.15)* | 0.99 (0.84,1.17)* |
| Spring | 0.94 (0.84,1.06)* | 0.83 (0.70,0.99)* | 1.01 (0.86,1.19)* |
| Year of birth | 0.95 (0.93,0.98)* | 0.97 (0.95,0.99)* | 1.02 (0.98,1.06)* |
| **Infant characteristics** |  |  |  |
| First Nations baby | 2.44 (2.15,2.77)* | 4.31 (3.53,5.24)* | 32.17 (19.76,52.36)* |
| Female | 0.91 (0.84,0.98) | 0.96 (0.85,1.08) | 0.93 (0.83,1.05) |
| Small for gestational age | 1.13 (0.99,1.28) | 1.01 (0.84,1.21) | 1.20 (1.01,1.43) |
| **Pseudo R<sup>2</sup> (aRR1)** | 0.15 | 0.16 | 0.26 |
| **Pseudo R<sup>2</sup> (aRR2)** | 0.15 | 0.16 | 0.26 |

**Supplementary Table 7.** Annual hospitalisation rates among Northern Territory infants in the first year of life (2008-2017): any code recorded during admission.

| **Any diagnosis**<br>Incidence rate (episode count) | **2008** | **2009** | **2010** | **2011** | **2012** | **2013** | **2014** | **2015** | **2016** | **2017** | **Overall** | **Annual trend**<br>**IRR (95% CI)** |
| --- | --- | --- | --- | --- | --- | --- | --- | --- | --- | --- | --- | --- |
| RSV-ALRI | 2.2 (66) | 3.4 (130) | 2.3 (87) | 2.7 (105) | 3.1 (123) | 2.0 (81) | 2.7 (105) | 2.1 (82) | 1.2 (46) | 1.5 (56) | 2.3 (815) | 0.91 (0.86,0.95) |
| ALRI | 9.9 (282) | 15.4 (586) | 16.1 (621) | 15.2 (591) | 12.6 (505) | 11.8 (468) | 11.2 (444) | 14.6 (584) | 11.2 (444) | 17.2 (667) | 13.9 (4910) | 0.99 (0.95,1.03) |
| *Viral pneumonia* | 0.5 (16) | 1.9 (71) | 1.3 (49) | 2.0 (76) | 1.5 (59) | 1.0 (39) | 1.3 (53) | 0.9 (36) | 0.8 (30) | 1.3 (49) | 1.3 (462) | 0.93 (0.87,0.98) |
| *Bacterial pneumonia* | 2.1 (62) | 3.6 (136) | 3.7 (142) | 3.9 (153) | 2.8 (111) | 2.2 (86) | 1.8 (69) | 2.6 (105) | 2.1 (85) | 3.0 (117) | 2.9 (1004) | 0.94 (0.90,0.99) |
| *Bronchiolitis* | 7.7 (217) | 10.8 (414) | 11.7 (454) | 11.3 (438) | 9.2 (368) | 9.2 (365) | 8.2 (323) | 11.2 (447) | 8.5 (338) | 12.2 (473) | 10.3 (3620) | 0.99 (0.96,1.03) |
| *Bronchitis* | 0.2 (8) | 0.1 (5) | 0.0 (1) | 0.1 (2) | 0 (0) | 0.1 (0) | 0.0 (1) | 0 (0) | 0.1 (2) | 0.2 (6) | 0.1 (19) | 1.03 (0.74,1.42) |
| *Non-specific ALRI* | 0.4 (12) | 1.3 (51) | 1.6 (61) | 1.1 (43) | 1.4 (55) | 0.7 (29) | 1.1 (43) | 1.7 (67) | 1.3 (50) | 2.7 (104) | 1.4 (503) | 1.06 (0.97,1.15) |

Annual rate data are presented as episodes per 100 child-years (episode count). Negative binomial regression was used to determine the annual trend in incidence rate with data presented as a crude incidence rate ratio (**IRR**); the effects of lead in truncation bias were avoided (for this birth cohort) by omitting data from 2008. **CI**: Confidence interval. **RSV-ALRI**: Respiratory syncytial virus specific acute lower respiratory infection.

## Geography classifications

Local Government Area boundaries were used to define Top End and Central desert regions (Australian Bureau of Statistics, https://maps.abs.gov.au/). ARIA+ (the Accessibility/Remoteness Index of Australia) classifications were used to define remoteness areas (https://able.adelaide.edu.au/housing-research/data-gateway/aria). There are three ARIA+ classifications in the NT: outer regional (Darwin and Palmerston region), remote (Greater Darwin and Alice Springs regions) and very remote (all other areas). Four geographical categories were used: (i) Top End, regional or remote; (ii) Top End, very remote; (iii) Central desert, remote; and (iv) Central desert, very remote.

## Notes

**Conflicts of Interest Disclosures:** We have no conflicts of interest to disclose.

**Funding/Support:** No funding was secured for this study.

### Competing Interest Statement

The authors have declared no competing interest.

### Funding Statement

No funding was secured for this study.

### Author Declarations

The project was approved by the Northern Territory Human Research Ethics Committee (2019-3551, 2018-3261) and Aboriginal Ethics Subcommittee. Individual-level consent was waived for this deidentified data. The project was also approved by The project was approved by the NT Human Research Ethics Committee (2019-3551, 2018-3261) and Aboriginal Ethics Subcommittee. Individual-level consent was waived for this deidentified data. The project was also approved by the Australian First Nations Reference Group for Child and Maternal Health at Menzies School of Health Research and the First Nations Advisory Group of the and Child and Youth Development Research Partnership.

